# Comparing methods to predict baseline mortality for excess mortality calculations

**DOI:** 10.1101/2022.07.18.22277746

**Authors:** Tamás Ferenci

## Abstract

**Background:** The World Health Organization (WHO)’s excess mortality estimates presented in May 2022 stirred controversy, due in part to the high estimate provided for Germany, which was later attributed to the spline model used. This paper aims to reproduce the problem using synthetic datasets, thus allowing the investigation of its sensitivity to parameters, both of the mortality curve and of the used method, thereby shedding light on the conditions that gave rise to this error and identifying possible remedies.

**Methods:** A negative binomial model was used accounting for long-term change, seasonality, flu seasons, and heat waves. Simulated mortality curves from this model were then analysed using simple methods (mean, linear trend), the WHO method, and the method of Acosta and Irizarry.

**Results:** The performance of the WHO’s method with its original parametrization was indeed very poor, however it can be profoundly improved by a better choice of parameters. The Acosta–Irizarry method outperformed the WHO method despite being also based on splines, but it was also dependent on its parameters. Linear extrapolation could produce very good results, but was highly dependent on the choice of the starting year, while the average was the worst in almost all cases.

**Conclusions:** Splines are not inherently unsuitable for predicting baseline mortality, but caution should be taken. In particular, the results suggest that the key issue is that the splines should not be too flexible to avoid overfitting. Even after having investigated a limited number of scenarios, the results suggest that there is not a single method that outperforms the others in all situations. As the WHO method on the German data illustrates, whatever method is chosen, it remains important to visualize the data, the fit, and the predictions before trusting any result. It will be interesting to see whether further research including other scenarios will come to similar conclusions.

## Introduction

Excess mortality is the difference between the actual all-cause mortality (number of deaths) over a particular time period in a given country or (sub- or supranational) region and its ‘‘expected” mortality, which refers to the mortality statistically forecasted from the region’s historical data. Excess mortality calculations can be used to characterize the impact of an event on mortality if the data were obtained before the onset of the event. Therefore, the prediction pertains to a counterfactual mortality that would have been observed without the event [1]. Excess mortality can thus measure the impact of the event, assuming that the prediction is correct.

Calculating excess mortality is particularly useful if the event’s impact on mortality is difficult to measure directly. For instance, one of the typical applications is characterizing the mortality associated with natural disasters [2–4], but it is also used for epidemics, such as the seasonal flu, where direct mortality registration is missing, incomplete, or unreliable [5,6].

Coronavirus disease 2019 (COVID-19) is no exception. While the mortality attributed to COVID-19 is reported in developed countries, typically daily or weekly, two drawbacks in the reporting have been realized. First, the number of reported deaths is – while to much less extent than the number of reported cases – still contingent on testing activity, which may be vastly different between countries or time periods. Second, despite efforts at standardization, the criteria for death certification may be different between countries [7]. Excess mortality resolves both of these problems because it is completely exempt from differences in testing intensity and cause-of-death certification procedures. This makes it especially suitable for between-country comparisons, which are critical to better understand the pandemic, particularly with regard to evaluating different control measures and responses [8].

This, however, comes at a price. First, and perhaps most importantly, excess mortality is inherently a gross metric, measuring both direct and indirect effects; the latter of which can be both positive (e.g., COVID-19 control measures also protect against the flu) and negative (e.g., the treatment of other diseases becomes less efficient) [9]. Second, excess mortality is the slowest indicator. The necessary data (i.e., the number of deaths) usually becomes available after 4 weeks (and even that is typically revised to some extent later) even in developed countries. This is in contrast to reported COVID-19 deaths, which are available by the following week or even the next day. Finally, the whole calculation depends on how accurate the forecast was.

The last of these issues is the focus of the current study. Given the importance of cross-country comparisons, the results should reflect true differences and should not be too sensitive to the prediction method used.

Only those methods that use the traditional regression approach are considered here. Methods using ARIMA models [10–13], the Holt-Winters method [14] or those based on Gaussian process [15] are not considered, nor are ensemble methods [16,17]. Questions concerning age or sex stratification or standardization [18], small area estimation [19,20], and inclusion of covariates (e.g., temperature) to improve modelling [16,17,19] are also not considered.

These considerations leave us with two matters of concern: the handling of seasonality and the handling of a long-term trend. For the latter, the following are the typical solutions concerning COVID-19:

- Using the last prepandemic year [16,21]. This solution is good – even if not perfect – because it uses data closest to the investigated period. However, this metric has a huge variance due to the natural year-to-year variability in mortality.
- Using the average of a few prepandemic years (typically 5 years) [22–28]. This is more reliable than the previous solution because averaging reduces variability. However, it is even more biased in case the mortality has a long-term trend (which it almost always has). For instance, if mortality is falling, this provides an overestimation; thus, excess mortality is underestimated.
- Using a linear trend extrapolation [29–31]. This approach accounts for the potential trends in mortality, removing the bias of the above-mentioned methods, at least as far as linearity is acceptable, but it depends on the selection of the starting year from which the linear curve is fitted to the data.
- Using splines [32,33]. The method of Acosta and Irizarry [34,35] is based on splines, just as many other custom implementations [16,36], which, crucially, includes the model used by the World Health Organization (WHO) [37].

While this issue has received minimal public attention, the choice of the method (and the parameters) used to handle long-term trends may have a highly relevant impact on the results of the calculation. This is evidenced by the case of excess mortality estimation by the WHO. On May 5, 2022, the WHO published its excess mortality estimates [38], which immediately raised several questions. In particular, it was noted that the estimates for Germany were surprisingly high [39]: the WHO estimated that 195,000 cumulative excess deaths occurred in Germany in 2020 and 2021, a figure that was inexplicably larger than every other previous estimate. For instance, the World Mortality Dataset reported 85,123 excess deaths in Germany for the same period [1].

This case was so intriguing, that one paper termed it as the “German puzzle” [39]. Figure 1 illustrates the “German puzzle” using actual German data, with the curves fitted on the 2015–2019 data and extrapolated to 2020 and 2021 (as done by the WHO). While the dots visually indicate a rather clear simple upward trend (as shown by the linear extrapolation), the spline prediction turns back.

**Figure 1:**
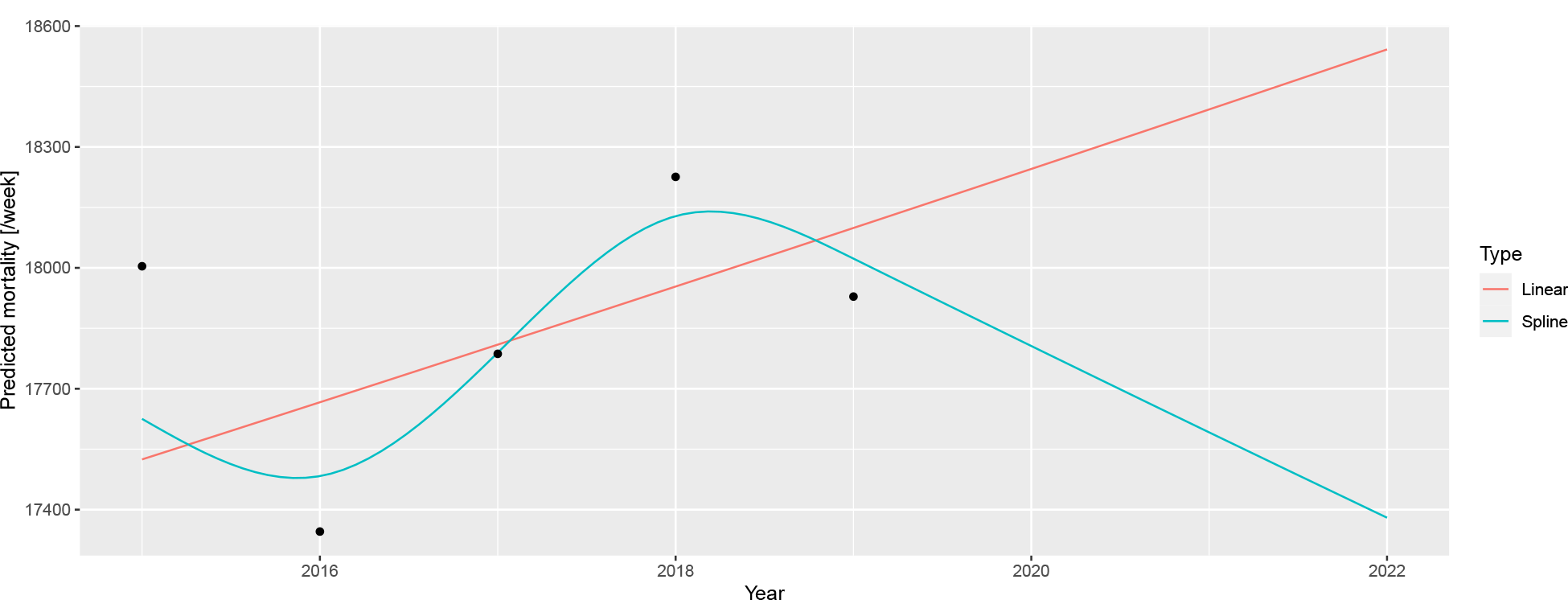
A linear trend and a spline fitted on the 2015–2019 German mortality data and extrapolated to 2020 and 2021.

The WHO [37] later explained that the problem resulted from two issues. First, the WHO rescaled the raw data to compensate for underreporting (caused by late registration, for instance), but this approach was unnecessary in the case of Germany, which had excellent death registration. Figure 1 shows the unadjusted German data avoiding this problem to focus on the second issue; that is, the usage of splines, which is the subject of the current investigation.

As described above, the WHO method uses a spline to capture the long-term trend. However, the lower data of 2019 had a very high impact on the spline, and this single observation turned the entire spline despite earlier points showing an upward trend. Too much weight seems to have been placed on this – likely short-term, random, noise-like – fluctuation; hence, the extrapolation was too sensitive for this. The culprit was quickly identified as the spline regression itself, with one commentator saying that “[e]xtrapolating a spline is a known bad practice” [39].

However, questions have emerged as to whether splines are really to be blamed. Motivated by the intention to better understand the ‘‘German puzzle,” this study aims to investigate the following questions: 1) Are splines really the culprit *per se*?, 2) What were the particular characteristics, both of the scenario and of the used spline regression used that gave rise to the problem?, and 3) Is there a better way to predict the baseline for the excess mortality calculation to avoid this problem?

To answer these questions, we first needed to devise a model that could generate mortality curves that capture the relevant features exhibited by the real-life German example. Thus, calculating the accuracy of a forecast would be possible because the ground truth is now known, and we could investigate how the parameters of the simulation influence it. By averaging several simulations, the mean accuracy can be approximated, allowing for a comparison of the methods and investigation of its dependence on the parameters – both for the mortality curve and for the method – thereby hopefully resolving the ‘‘German puzzle.”

This investigation focuses on the errors of prediction. However, as excess mortality is defined as the difference between actual and predicted mortality, any error in the prediction directly translates to the same error in the estimation of excess mortality (given any actual mortality). Thus, the current study equivalently covers the errors in the estimation of excess mortality itself.

It should be noted that the present study does not use age or sex stratification or consider the size and composition of the background population. While these parameters are important, the WHO’s original study also did not take these factors into account.

## Methods

### Data source

Data on weekly all-cause mortalities for Germany were obtained from the European Statistical Office (Eurostat) database demo_r_mwk_ts. We applied no additional pre-processing or correction such as that for late registration, as part of the problem with the WHO’s approach was caused by upscaling, and we wanted to focus solely on the modelling aspect. Additional File 1 shows a detailed comparison of the possible data sources.

Figure 2 illustrates the basic properties of the data (raw weekly values, yearly trend, and seasonal pattern).

**Figure 2:**
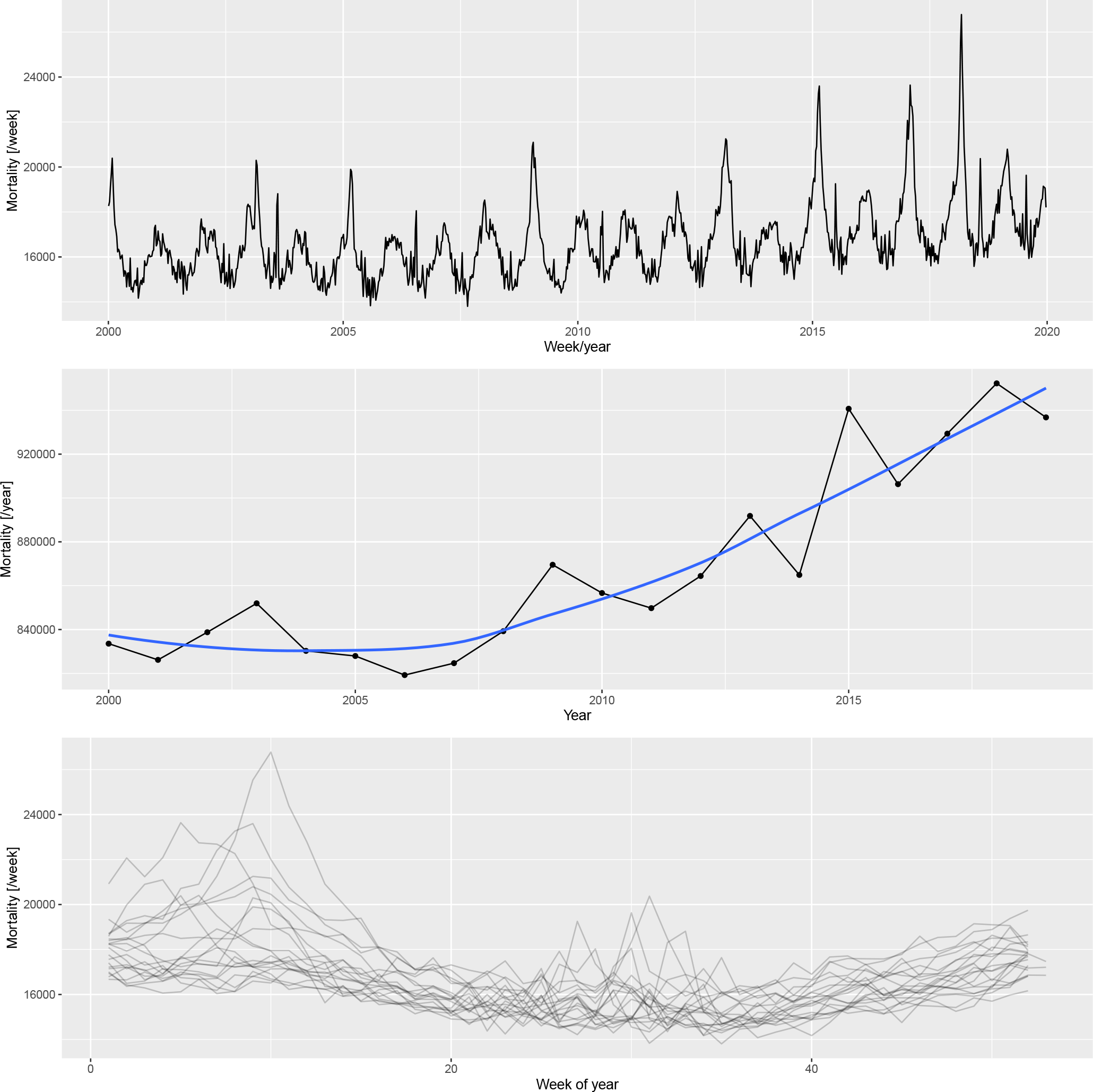
Weekly mortalities (upper), yearly mortalities with the LOESS-smoother (middle), and the seasonal pattern (bottom) for the German mortality data, 2000–2019.

### Simulation model

Based on the patterns shown in 2, the following three components were used to create the synthetic datasets:

- Long-term change, modelled with a quadratic trend; described by three parameters (constant, linear, and quadratic terms)
- Deterministic seasonality, modelled with a single harmonic (sinusoidal) term; described by two parameters (amplitude and phase)
- Random additional peaks during winter (i.e., flu season) and summer (i.e., heat waves); described in each season by five parameters (probability of the peak, minimum and maximum values of the peak height, and minimum and maximum values of the peak width)

These components governed the expected value; the actual counts were obtained from a negative binomial distribution with a constant size and log link function.

Thus, the number of deaths at time *t* (*D*_*t*_), was simulated according to a negative binomial model *D*_*t*_ ∼ NegBin (*μ*_*t*_, *s*), with mean *μ*_*t*_ and size parameter *s* = 1000. The mean was modelled such that log (*μ*_*t*_) = *β*_0_ + *β*_1_ · *t* + *β*_2_ · *t*^2^ + *A* · cos (2*π* · *w* [*t*] + *φ*), where *w* [*t*] is the week of time *t*, scaled from 0 to 1. The first three terms in this equation refer to the “long-term trend” (characterized by parameters *β*_0_ = 10.11, *β*_1_ = −7.36 · 10^*−*5^, and *β*_2_ = 3.04 · 10^*−*9^), and the latter to the seasonality (characterized by parameters *A* = 0.07 and *φ* = −0.61). Additionally some peaks have been randomly added to log (*μ*_*t*_), with widths between 8.41 and 35.36 and heights between 0.11 and 0.33 positioned in the winter (uniformly between weeks 1 and 11) with 0.45 probability in each year, and with widths between 0.86 and 9.24 and heights between 0.10 and 0.24 positioned in the summer (uniformly between weeks 26 and 37) with 0.40 probability in each year. The shape of the peaks follows the probability density function of the Cauchy distribution. These parameters were chosen so that the simulated curves mimic the properties of the real-life mortality curve.

Additional File 2 describes in detail how the above-mentioned model is built (including the estimation from the actual German data that resulted in these parameters, and an example of the simulated data along with the real data).

### Baseline mortality prediction

We predicted mortality by using four methods: the WHO method, an advanced alternative method developed by Acosta and Irizarry in 2020 that also uses splines [34], and two simple methods for comparison. These latter methods cover classical statistical methods that are widely used for predicting baseline mortality in excess mortality studies. The methods are detailed below:

- Average: after accounting for seasonality with a single cyclic cubic regression spline, the average of the preceding years was used as the constant, predicted value. The response distribution is assumed to be negative binomial (with the overdispersion parameter estimated from the data) with a log link function. Parameter: starting year (i.e., how many previous years were used for averaging). Some studies used the last prepandemic year (2019) as the predicted baseline mortality, this is just the special case of this method, with the starting year set to 2019.
- Linear: after accounting for seasonality with a single cyclic cubic regression spline, the long-term trend was modelled with a linear trend. The response distribution is assumed to be negative binomial (with the overdispersion parameter estimated from the data) with a log link function. Parameter: starting year (from which the model was fitted).
- WHO method: this method was reconstructed according to the description mentioned above [37]. Briefly, seasonality was accounted for with a single cyclic cubic regression spline (as done in previous cases), and the long-term trend was accounted for with a thin plate regression spline. The only deviation compared with WHO’s study is that the actual time (number of days since January 1, 1970) was used as the predictor of the long-term trend, not the abruptly changing year. The response distribution is assumed to be negative binomial (with the overdispersion parameter estimated from the data) with a log link function, and the model was estimated with restricted maximum likelihood. Second derivative penalty was used for constructing the spline; thus, the forecasting would be a linear extrapolation. Parameters: starting year (from which the model is fitted) and *k*, which is the dimension of the basis of the spline used for capturing the long-term trend.
- Acosta–Irizarry (AI) method: the method described in [34] using their reference implementation. It offers many advantages when estimating excess mortality, however, these advantages partly appear only in the second stage (i.e., calculating the excess after the expected is forecasted). Regarding baseline prediction, the method is similar to that of the WHO in using splines, with three differences. First, for capturing seasonality, two harmonic terms are used (with prespecified frequencies of 1/365 and 2/365 as default, and arbitrary phase estimated from the data) instead of the cyclic cubic regression spline. Second, the spline to capture the long-term trend is a natural cubic spline, not a thin plate regression spline, with the number of knots selectable. If the number of years in the training data is less than 7, a linear trend is used instead of the spline. Finally, the response distribution is quasi-Poisson (with a log link function). Parameters: starting year (from which the model is fitted) and *tkpy*, which denotes the number of trend knots per year. Other parameters are left on their default values (i.e., two harmonic terms are used).

The equations presented in Table 1 provide an overview of these modelling approaches.

**Table 1.** Overview of the models used to create predictions. cc: cyclic cubic regression spline, tp: thin plate regression spline, nc: natural cubic spline, *: >7 years training data, **: ≤ 7 years training data, *w* [*t*]: week of the year scaled to 0-1.

| Name | Model |
| --- | --- |
| Average | $Y_t \sim NegBin(\mu_t, \theta) \log(\mu_t) = \beta_0 + f_{cc}(w[t])$ |
| Linear | $Y_t \sim NegBin(\mu_t, \theta) \log(\mu_t) = \beta_0 + \beta_1 t + f_{cc}(w[t])$ |
| WHO | $Y_t \sim NegBin(\mu_t, \theta) \log(\mu_t) = f_{tp}(t) + f_{cc}(w[t])$ |
| Acosta-Irizarry | $Y_t \sim QuasiPoi(\mu_t) \log(\mu_t) =$<br>$\begin{cases} \beta_0 + f_{nc}(t) + \sum_{k=1}^2 [\sin(2\pi \cdot k \cdot w[t]) + \cos(2\pi \cdot k \cdot w[t])] & * \\ \beta_0 + \beta_1 t + \sum_{k=1}^2 [\sin(2\pi \cdot k \cdot w[t]) + \cos(2\pi \cdot k \cdot w[t])] & ** \end{cases}$ |

As already noted, population size is not included in the models, consistent with what the WHO did for the analysis of countries with frequently available data. Thus, changes in the population size are absorbed into the changes in death counts without explicit modelling, clearly suggesting potential room for improvement.

### Validation through simulation

First, a synthetic dataset was randomly generated from the model described above using the investigated parameters (parameters of the scenario). This dataset simulated mortalities recorded from 2000 to 2023. We then applied the investigated prediction method with the investigated parametrization (parameters of the method), and after fitting on the 2000–2019 data, we obtained a prediction for 2020–2023, where it can be contrasted with the simulation’s actual values, which represent the ground truth in this case. Denoting the actual number of deaths in the simulated dataset for year *y* with *M*_*y*_ = Σ_*t*:*y*[*t*]=*y*_ *D*_*t*_ and the predicted number with 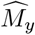, the goodness of prediction is quantified with mean squared error 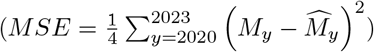, mean absolute percentage error 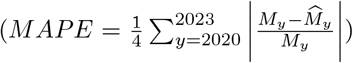, and bias 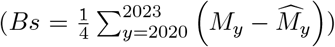. This whole procedure was repeated 1000 times, and the metrics were averaged over these replications. This step was then repeated for all prediction methods, all parameters of the method, and all parameters of the scenario.

The investigated parameters of the prediction methods were the following:

- Average: starting years 2000, 2005, 2010, 2015, 2019
- Linear: starting years 2000, 2005, 2010, 2015
- WHO method: all possible combination of starting years 2000, 2005, 2010, 2015 and *k* (basis dimension) 5, 10, 15, 20
- AI method: all possible combination of starting years 2000, 2005, 2010, 2015 and *tkpy* (trend knots per year) 1/4, 1/5, 1/7, 1/9, 1/12

For the scenario, simulations were run with the optimal parameters to mimic the real-life German situation, as discussed previously (base case scenario), and three further parameter sets, describing a scenario where the long-term trend is linear (*β*_0_ = 10.11, *β*_1_ = −7.36 · 10^*−*5^ and *β*_2_ = 0), constant (*β*_0_ = 10.11, *β*_1_ = 0 and *β*_2_ = 0), and when it is non-monotone (*β*_0_ = 10, *β*_1_ = 9.5 · 10^*−*5^ and *β*_2_ = −3 · 10^*−*9^).

This framework also enables us to investigate any further scenarios, including varying parameters other than the long-term trend, or varying several in a combinatorial fashion (although the latter has a very high computational burden).

Additional File 3 details the simulation.

### Programs used

All calculations were performed using the R statistical program package version 4.3.1 [40] with the packages `data.table` [41] (version 1.14.8), `ggplot2` [42] (version 3.4.3), `excessmort` (version 0.6.1), `mgcv` (version 1.8.42), `scorepeak` (version 0.1.2), `parallel` (version 4.3.1) `lubridate` (version 1.9.3), `ISOweek` (version 0.6.2) and `eurostat` (version 3.8.2).

The full source code allowing for complete reproducibility is openly available at: https://github.com/tamas-ferenci/MortalityPrediction.

## Results

Figure 3 illustrates the 2020–2023 predictions for the base case scenario by showing the estimated yearly deaths for 200 randomly selected simulations together with the ground truth for all four methods with all possible parameters.

**Figure 3:**
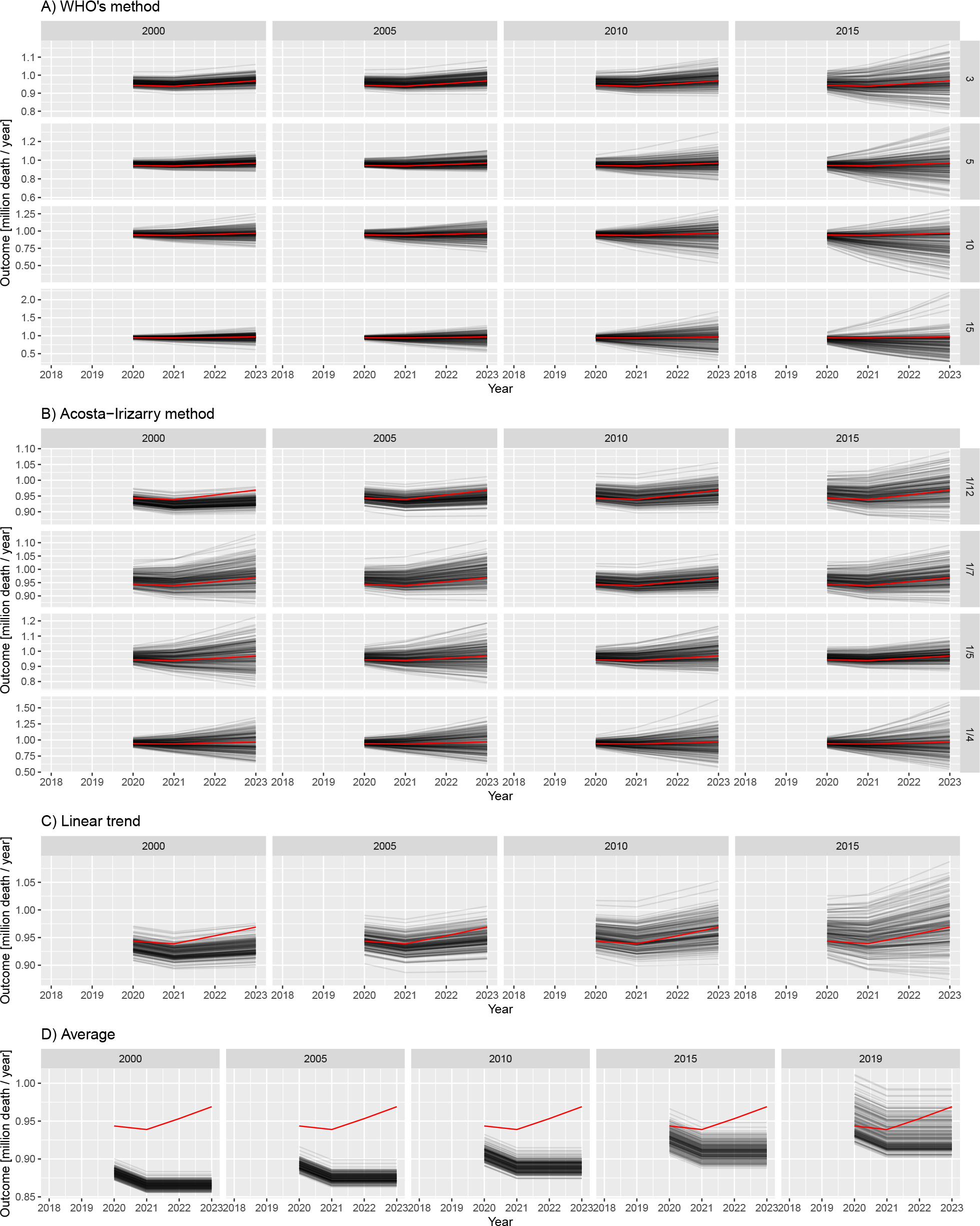
Estimated yearly deaths (for 2020–2023) for 200 randomly selected simulations (black lines) together with the ground truth (red line). A) WHO method, B) Acosta–Irizarry method, C) Linear trend, D) Average. Parameters of the methods are shown in column and row headers, and parameters of the scenario are set to the base case values. Note that 2020 is a long year with 53 weeks; therefore, higher values are expected for that year.

Figure 3 already strongly suggests some tendencies but, for a precise evaluation, we needed to calculate the error metrics. Figure 4 shows the three error metrics for all methods and all possible parametrizations. As shown in the figure, the ordering of the methods according to different criteria is largely consistent.

**Figure 4:**
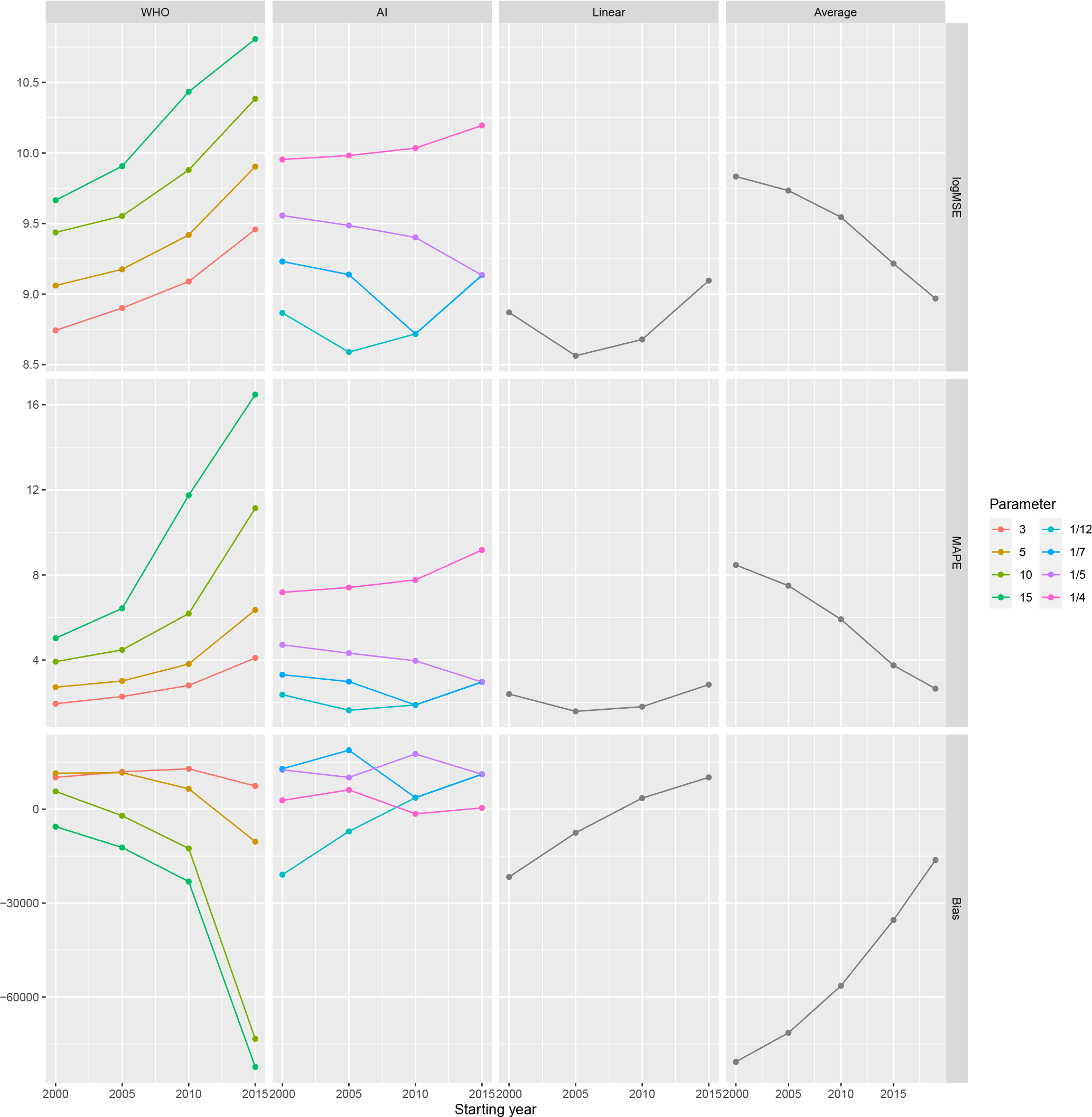
Different error metrics (logMSE, MAPE, and Bias) for all methods and all possible parameter combinations of all methods. Parameters of the scenario are set to the base case values.

As suggested in Figure 3, this confirms that *k* = 3 (WHO) and *tkpy* = 1*/*12 or 1*/*7 (AI) are the best parameters in this particular scenario. Note that the default value in the method used by the WHO is *k* = 10, but it is just *tkpy* = 1*/*7 for the AI method.

Figures 3 and 4 shed light on the nature of error. The linear trend and average methods are particularly clear in this respect: early starting ensured low variance, but it was highly biased. Conversely, later starting reduced the bias but increased the variance. Thus, this observation is a typical example of a bias-variance trade-off.

All the above-mentioned investigations used the base case scenario for the simulated mortality curve. Figure 5 shows the MSEs achievable with each method in the further investigated scenarios representing four distinct types of the long-term trend of simulated mortality as a function of the starting year (with *k* = 3 for the WHO method and *tkpy* = 1*/*7 for the AI approach).

**Figure 5:**
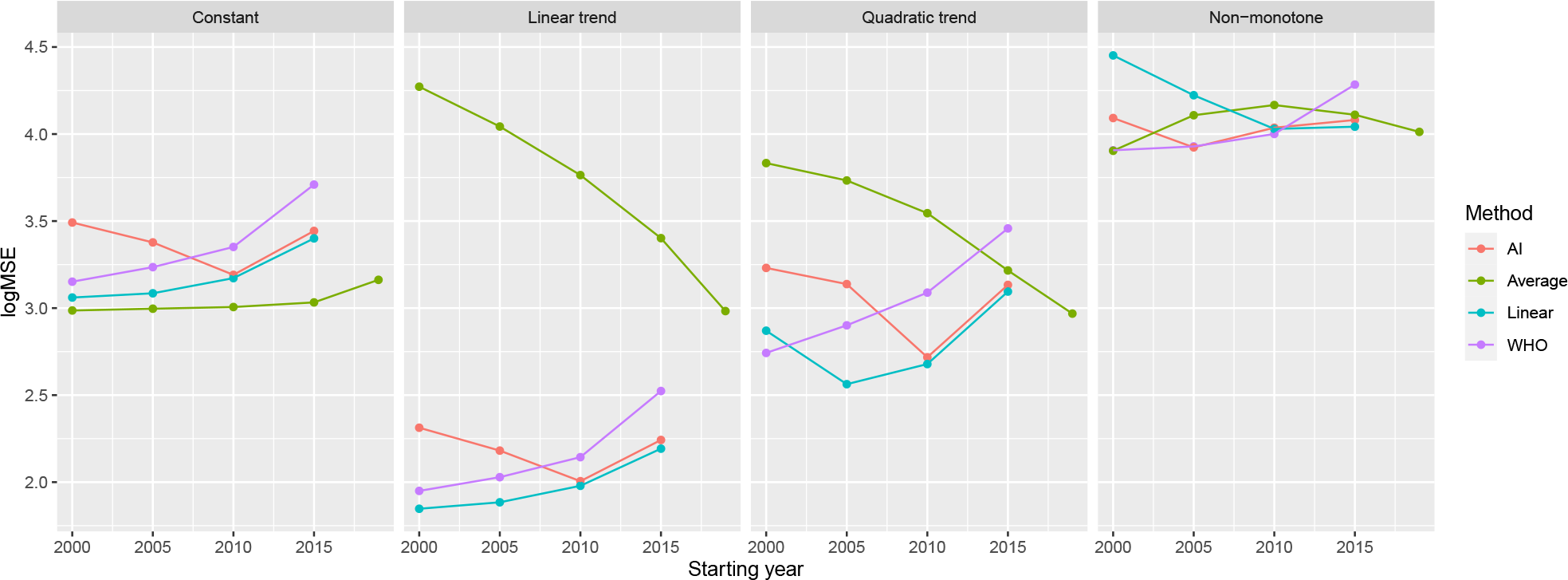
Mean squared errors of the investigated methods by starting year on a logarithmic scale (with k = 3 for the WHO method and tkpy = 1/7 for the AI approach) for the four defined scenarios.

Table 2 summarizes the error metrics of all four methods.

**Table 2.** Mean squared errors of the investigated methods, using the parametrization with the lowest MSE for each scenario (data shown as mean ± standard deviation).

| Method | Constant | Linear trend | Quadratic trend | Non-monotone |
| --- | --- | --- | --- | --- |
| AI | 1182.2 ± 1273.6 | 73.3 ± 77.4 | 388.2 ± 553.5 | 8380.2 ± 14478 |
| Average | 969 ± 596 | 961.3 ± 753.1 | 929.6 ± 1039.6 | 8013.8 ± 4529.7 |
| Linear | 1150 ± 1213.7 | 70.3 ± 73.1 | 365.5 ± 524 | 10702.6 ± 11901.7 |
| WHO | 1418.4 ± 2075.1 | 89.1 ± 123.8 | 552.5 ± 824.5 | 8067.2 ± 9866.4 |

Finally, considering that different methods were evaluated on the same simulated dataset for each simulation, we could directly compare not only the averages but also the errors themselves. Additional File 4 presents this possibility.

## Discussion

The results of this study demonstrate that we could reliably reproduce the “German puzzle” using synthetic datasets. This approach allowed us to investigate how the results depend on the method used, its parameters, and the parameters of the scenario.

As expected, prediction with averaging had the highest error, except for very poor parametrizations of the spline-based methods, and it was highly biased. Its performance was improved by a later starting year; that is, a smaller bias offsets the larger variability. Naturally, it performs well in a practically very unlikely case when even true mortality is constant.

Linear extrapolation seemed to be a very promising alternative, comparable to the considerably more sophisticated spline-based methods. The only problem was that it was very sensitive to the appropriate selection of the starting year; the phase where the change in historical mortality is linear should be covered. Linear extrapolation is prone to subjectivity and may not work at all if the linear phase is too short (thereby limiting the available information), but it depends on how wiggly the historical curve is. Naturally, it works best if the true mortality is linear, but it can perform very poorly with non-monotone curves when the starting year is not selected to match the last section that can be approximated with a linear curve, consistent with the previous remark.

Theoretically, splines can work well even in cases of non-monotone mortality. The spline-based method can use all historical data. It is not abruptly cut off as with linear extrapolation, but more weight is placed on the trends suggested by recent observations. At first glance, this method seems to be the ideal solution, delivering the benefits of linear extrapolation but without the need to ‘‘guess” the good starting point. However, as this study reveals, the definitions of ‘‘more weight” and ‘‘recent” are crucial, and certain parameter choices can result in very poor extrapolations, despite the tempting theoretical properties.

In selecting the optimal parameters, as confirmed by the results of the two spline-based methods (the WHO method and the AI method), the splines should be quite simple in baseline mortality prediction for excess mortality calculations. This finding is in line with the experiences both with the WHO method (where increasing the basis dimension decreased the performance) and the AI method (where increasing the trend knots per year decreased the performance).

One possible explanation is that mortality curves exhibit only slow changes, so high flexibility is not required. As with any regression model, excessively high model capacity can be downright detrimental, as it allows the model to pick up noise, which can cause overfitting.

Importantly, the AI method only uses splines to model the long-term trend if it has more than 7 years of data. Otherwise, it switches into a simple linear trend. This finding is completely in line with the above-mentioned remarks. That is, flexibility is useful but can backfire with small amounts of training data.

In Germany, the 2019 data were quite lower, likely because of simple random fluctuation, but the spline was flexible enough to be ‘‘able to take this bend.” Data are presented using the ISO 8601 year definition; thus, the year can be either 52 or 53 weeks long [43]. From 2015 to 2019, every year was 52 weeks long, except for 2015, which was 1 week longer. This definition adds to the reasons why the value for 2015 is higher, increasing the German data’s wiggliness and thereby potentially contributing to the problem. The increased wiggliness of the data forces the thin plate regression spline used in the WHO method to be more flexible. (This was not a problem with the WHO’s original analysis because it used monthly data, but it could be if the WHO method is directly applied to weekly data.)

The WHO method is only acceptable if *k* ≤ 5 (but even that requires a longer observation than starting from 2015, as was done by the WHO), not higher. For the AI method, 1/4 trend knots per year was too flexible, and perhaps even 1/5 was too high. The default value in the reference implementation of the AI method is 1/7, and using a considerably higher value is not recommended. However, the WHO study did not specify what basis dimension was used [37]; nonetheless, the default of the package used is *k* = 10. Thus, even *k* = 5, and especially *k* = 3, is substantially lower. Possibly, this component is crucial in the WHO’s experience, where the starting year was 2015 (and *k* = 10 was probably used).

Between the two spline-based methods, when using rigidity parameters that are optimal in this particular scenario, the WHO method performed better with longer fitting periods, whereas the AI method performed better with shorter ones. However, the performance of the AI method was considerably less dependent on the starting year.

Perhaps one of the most important lessons learned, especially from Figures 4 and 5, is that there is no ‘‘one size fits all” optimal choice. In other words, a method that performs well for a given true mortality curve can exhibit extremely poor performance for another shape of mortality. Moreover, the optimal choice of parameters even for one given method can substantially depend on the scenario, and a parameter that works well for one situation might be poor for another. Hence, while there are ‘‘safer choices,” recommending a universally ‘‘optimal” parameter is not possible. In selecting a good parameter for a particular case, perhaps the most important is to examine the fit of the model on historical data. Figure 6 provides an example using the German data. The figure shows the prediction for 3 years (2020–2022) from the historical data, using all methods and all parameters. A quick visual inspection immediately reveals relevant and likely meaningless predictions. The latter, unfortunately, includes that of the WHO (2015 as the starting year, *k* = 10). Thus, such an inspection would have likely revealed the problem. Time-series bootstrap and time-series cross-validation, which are objective, might be promising alternatives for the potentially subjective method of visual inspection, still with the use of only historical data.

**Figure 6:**
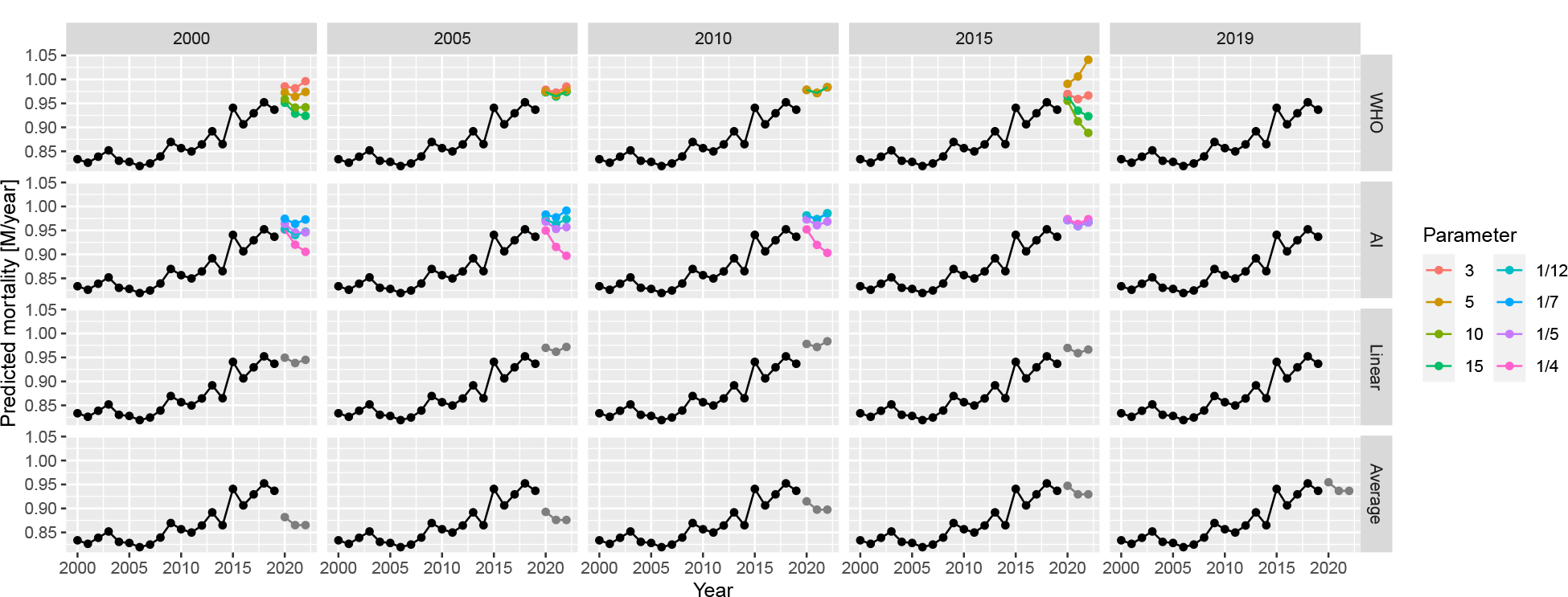
Predictions for 2020–2022 from all methods with all possible parameters, using historical German data.

Two studies conducted by Nepomuceno et al. [44] and Shöley [45] are similar to ours. They used several models, partly overlapping those that are presented in our study but, importantly, neither of them considered splines for a long-term trend. Nepomuceno et al. did not try to evaluate the methods and compared these methods without having ground truth to determine any concordance. In contrast, Shöley conducted an objective evaluation but, contrary to the synthetic dataset simulation approach used in the present study, time-series cross-validation with historical data was applied to measure accuracy. Cross-validation is guaranteed to be realistic as opposed to a simulation, but it has less freedom; in this method, investigators are bound to the empirical data with limited possibilities in varying the parameters.

We did not investigate the impact of using the population and modelling death rates instead of death counts directly. We also did not examine the potential impact of data frequency. In our study, we used weekly data, which may be unavailable in developing countries but are almost universally available in developed countries. Therefore, using the most frequent data seems logical (with the appropriate handling of seasonality). Furthermore, because our focus was on developed countries, we did not consider adjusting for late registration and imputing missing data, which may be needed when full data are unavailable.

Another limitation of our study is that it only analysed point estimates. The applied prediction models can provide prediction intervals; thus, investigating their validity (e.g., coverage properties) could be a relevant future research direction.

Finally, this study did not consider the age and sex structures of the population (consistent with the WHO’s approach). However, the inclusion of these structures together with an explicit modelling of the population size might improve predictions by capturing and separating mechanisms that govern the change of population size and structure in the models. To explore whether and to which extent predictions could be improved by taking these structures into account is an important matter that requires further research.

## Conclusion

The performance of the WHO’s method with its original parametrization is indeed very poor, as revealed by extensive simulations. Thus, the ‘‘German puzzle” was not just an unfortunate mishap, but it could have been profoundly improved by selecting better parameters. The performance of the WHO method is similar to that of the AI method, with the former being slightly better for longer fitting periods and the latter for shorter ones. Despite its simplicity, linear extrapolation can exhibit a favourable performance, but this depends highly on the choice of the starting year. Conversely, the AI method exhibits relatively stable performance (considerably more stable than the WHO method) irrespective of the starting year. The performance of the average method is almost always the worst, except for very special circumstances.

This study also shows that splines are not inherently unsuitable for predicting baseline mortality, but caution should be taken, with the key issue being that the splines should not be too flexible to avoid overfitting. In general, no single method outperformed the others in the investigated scenarios. Regardless of the approach or parametrization used, it is essential to have a proper look at the data, and to visualize the fit and the predictions produced by the method used. Further research is warranted to see if these statements can be confirmed on the basis of other scenarios.

## Data Availability

All data produced are available online at https://github.com/tamas-ferenci/MortalityPrediction.

## Additional File 1: Comparison of data sources

For a country like Germany, four data sources come into consideration for weekly mortality data: Eurostat [46], the Short-Term Mortality Fluctuations (STMF) dataset of the Human Mortality Database (WMD) [47], the World Mortality Database [1] and the national data provider (in this case, the Federal Statistical Office of Germany). The last is usually more complicated, limits extension to other countries and is unnecessary for developed countries, so it’ll be avoided in this case. Also, for Germany, WMD simply copies the data of the STMF (“We collect the weekly STMF data for the following countries: […] Germany, […].”) leaving us with two options.

We shall compare whether these two report identical data (Figure 7).

**Figure 7:**
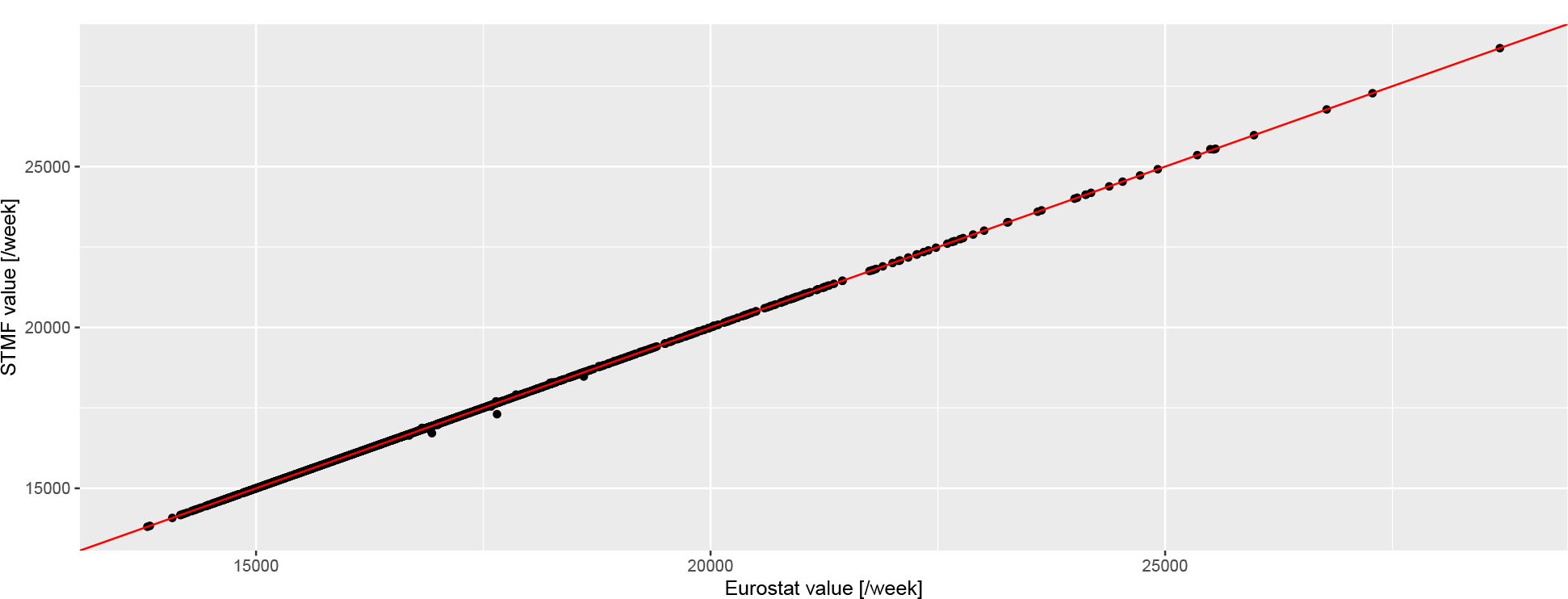
Weekly number of deaths according to the Eurostat (horizontal axis) and the STMF database (vertical axis) in Germany. Red line indicates the line of equality.

The two are almost identical (with a correlation of 0.9999781), with differences only occuring for the latest data and of minimal magnitude, so we can safely use the Eurostat database.

## Additional File 2: Generating realistic synthetic datasets

First, Figure 2 should be inspected, as it already gives important clues on the setup of a realistic model from which synthetic datasets could be generated. Figure 8 gives further insight by plotting each year separately.

**Figure 8:**
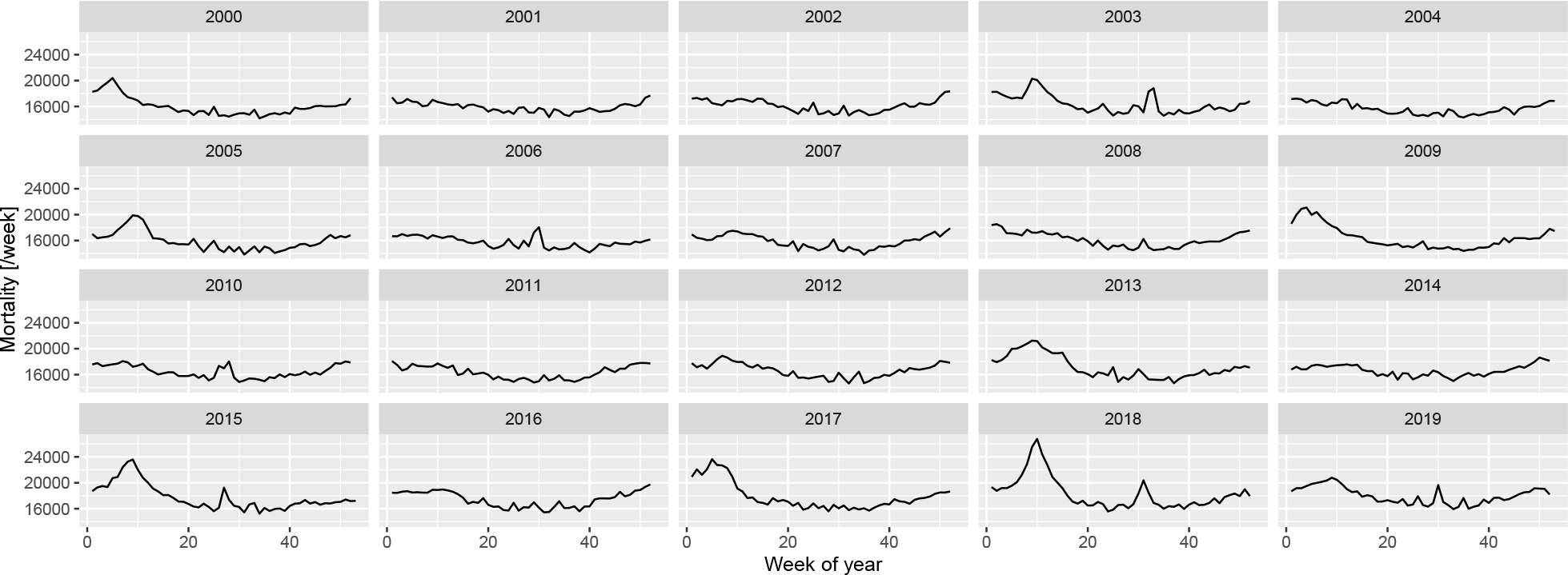
Weekly number of deaths in Germany, separated according to year.

The following observations can be made:

- There is a long-term trend, seemingly quadratic.
- There is a strong seasonality with winter peak and summer trough.
- There are peaks – in addition to the seasonality – in the winter and also during the summer (although the shape seems to be different, with winter peaks seeming to be broader and higher).

To investigate these, first a spline-smoothing – with thin plate regression spline [32] – is applied to obtain the long-term trend, and a single harmonic term is included as a covariate to remove seasonality. No interaction is assumed between the two, i.e., it is assumed that the seasonal pattern is the same every year. (The peaks are not accounted for at this stage which means that the curve is above the true one, but the difference is likely has minimal due to the rarity and short duration of the peaks. This will be later corrected, after the peaks were identified.) All analysis will be carried out on the log scale (meaning the effect of covariates is multiplicative) using negative binomial response distribution to allow for potential overdispersion [48].

Figure 9 shows the results, overplotted with the model where the long-term trend is a completely parametric quadratic trend. One can observe very good fit between the two, so all subsequent investigation will use the quadratic trend which is much easier to handle. This is only meaningful for short-term extrapolation, but this is what will be needed now (two years of extrapolation will be used in the present study); also it is not possible to better differentiate between functional forms at this sample size.

**Figure 9:**
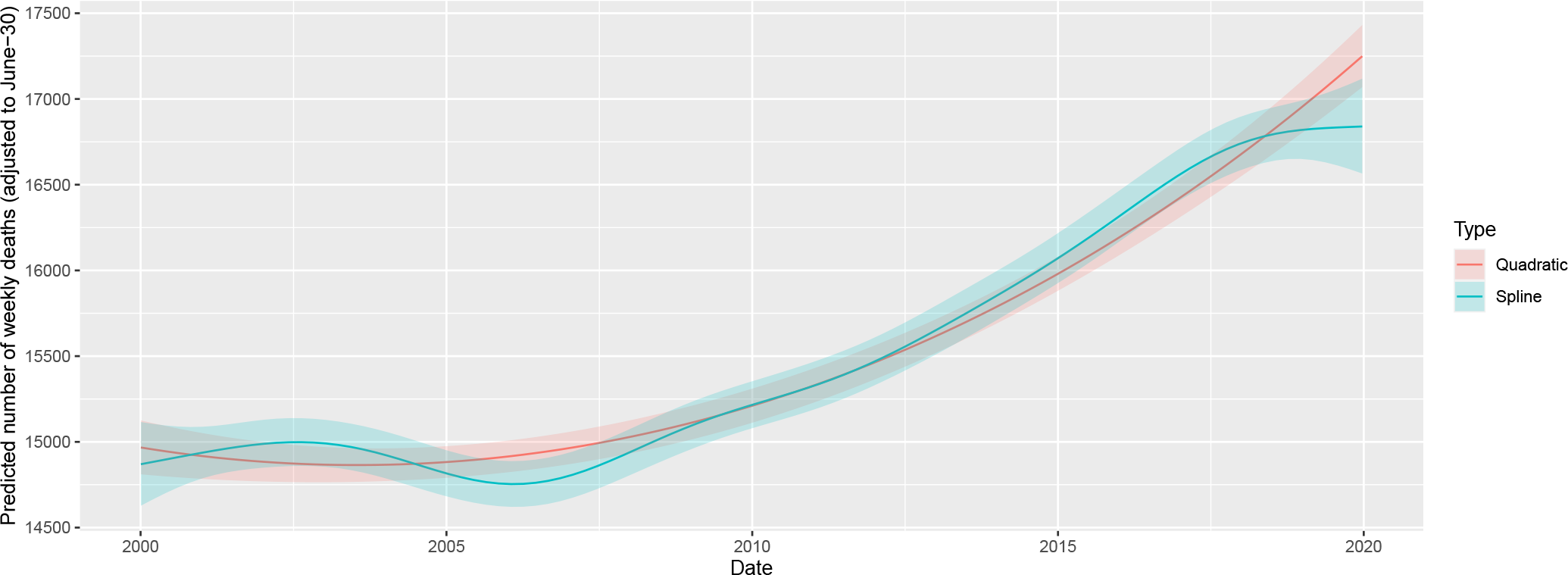
Fitting long-term trend as spline (black) and as quadratic trend (red); shaded area indicated 95% confidence interval. Seasonality is removed by including a single harmonic term in the regression in both cases.

The coefficients can be transformed to equivalent forms that are more meaningful. Thus, the three parameters of the quadratic trend can be expressed as a minimum point (2003-07-15), value at the minimum (15918.54) and value at the end of 2020 (18825.83). (This differs from the value seen on Figure 9, as that also includes the effect of the harmonic term.) The two parameters of the harmonic regression can be expressed as an amplitude, a multiplier (9.4%) and a phase shift (-0.7, i.e., minimum at week 32 of the year).

Figure 10 shows the predictions of the above model.

**Figure 10:**
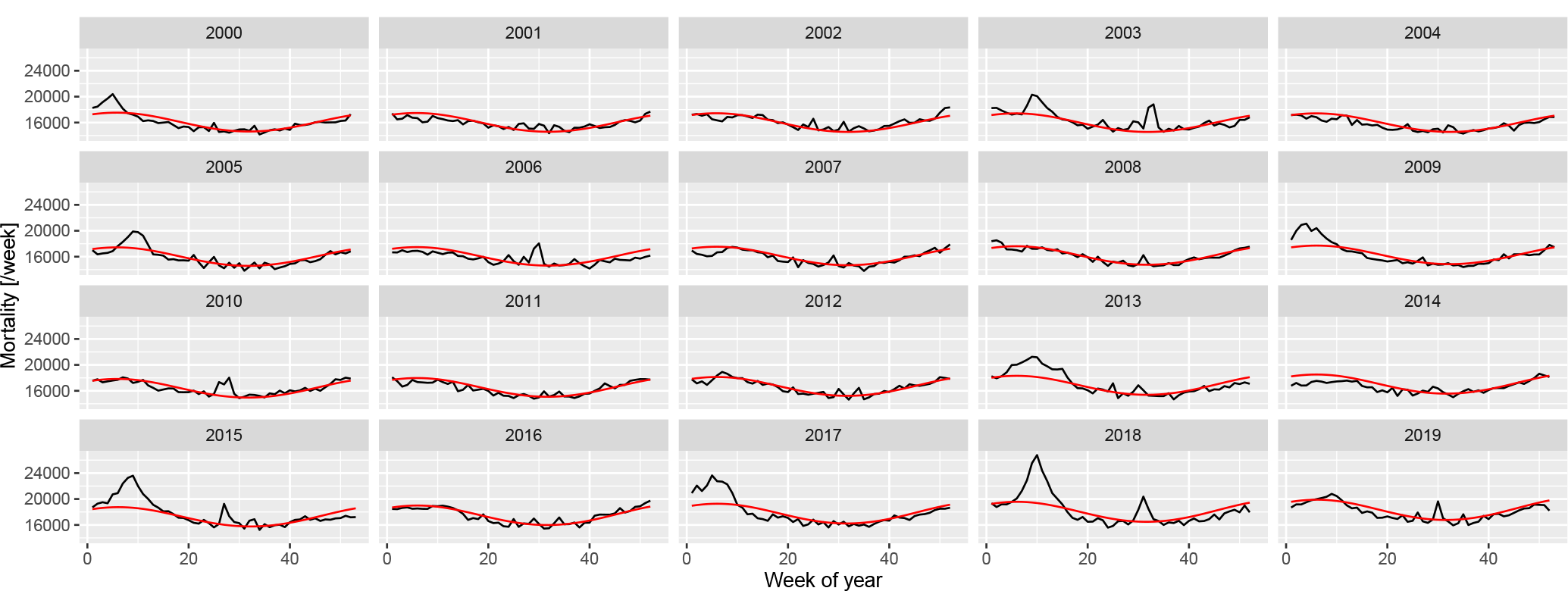
Weekly number of deaths in Germany, separated according to year, showing the predictions from the model with quadratic long-term trend and a single, fixed harmonic term.

A good fit can be observed, apart from summer and winter peaks. Thus, to capture them, the predictions are subtracted; with the results shown on Figure 11.

**Figure 11:**
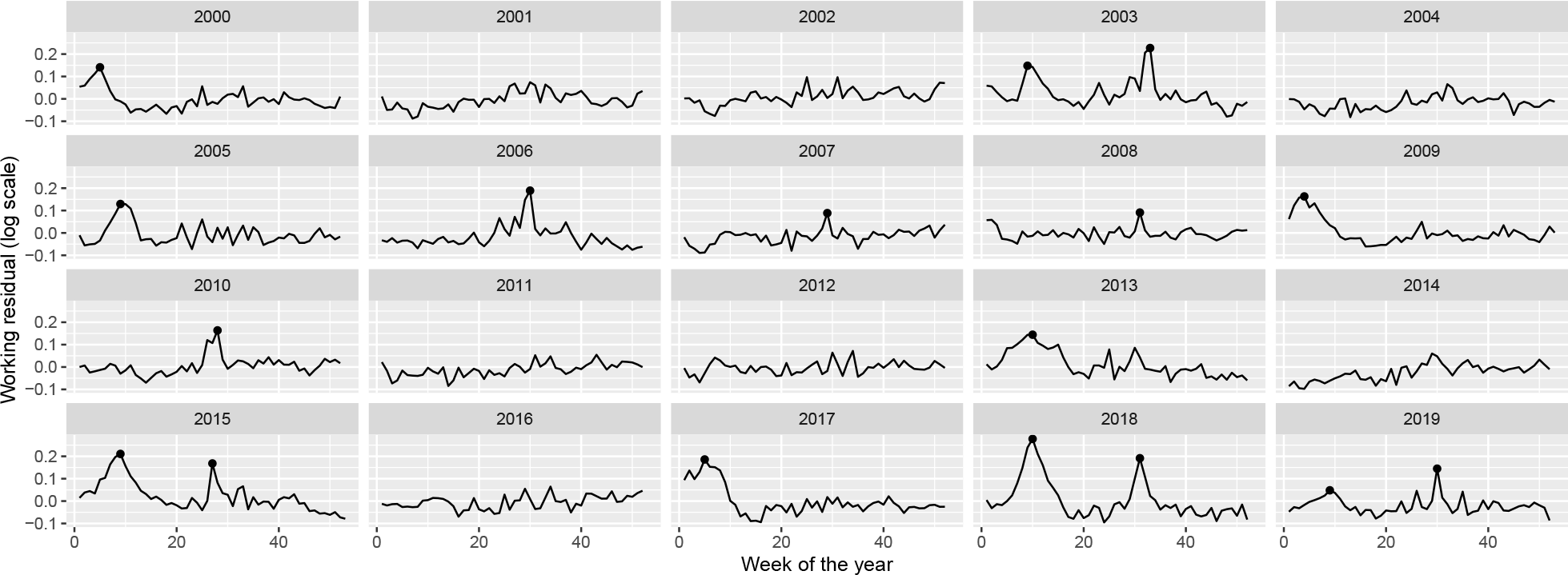
Residuals of the fitted model with quadratic long-term trend and a single, fixed harmonic term. Dots indicate identified peaks.

Peaks in the residuals were identified with the peak detector of Palshikar [49] using parameters that were empirically tuned to identify to visually clear peaks. Results are shown on Figure 11 with black dots; an indeed good identification of the unequivocal peaks can be seen.

Figure 12 shows the peaks themselves with a ± 100 days neighbourhood. This reinforces the idea that summer and winter peaks are somewhat different, but more importantly, suggests that the rescaled probability density function of the Cauchy distribution, i.e., 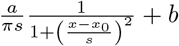 might be a good – and parsimonious – function form to capture the shape of the peaks.

**Figure 12:**
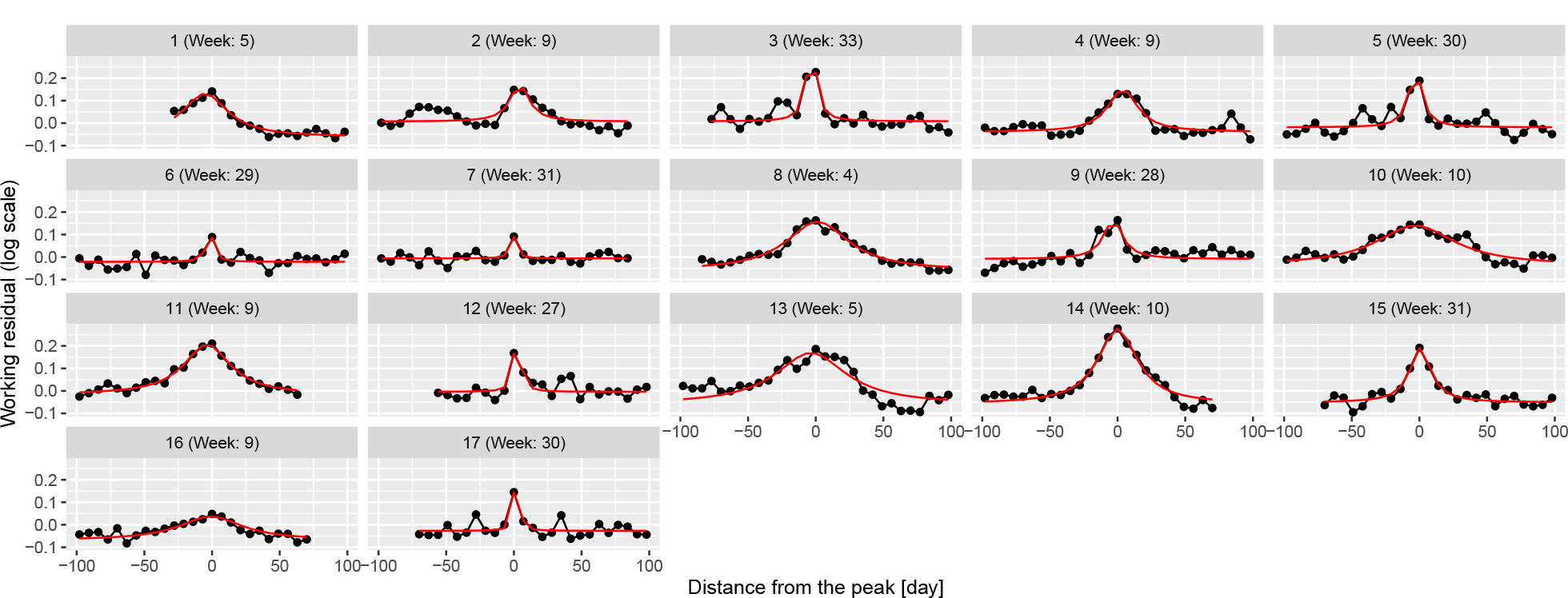
100-day width neighbourhood of the identified peaks. Red line indicates the best fitting rescaled Cauchy density.

To check this theory, the best fitting function was found for each peak individually using the Nelder-Mead method [50] with mean squared error objective function. Results are shown on 12 as red lines; an almost perfect fit can be observed for all peaks confirming the initial idea of using Cauchy density.

This now puts us in a position to investigate the distribution of the parameters (i.e., *a, b, s* and *x*_0_), which is shown on Figure 13 separated according to whether the peak is during the summer or not. Peak height is also calculated, defined as height at zero (which is 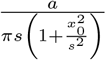) not the actual maximum height (which is at *x*_0_) to avoid extremely large heights – which were never actually observed – due to peaks with small *s*, i.e., very narrow peaks.

**Figure 13:**
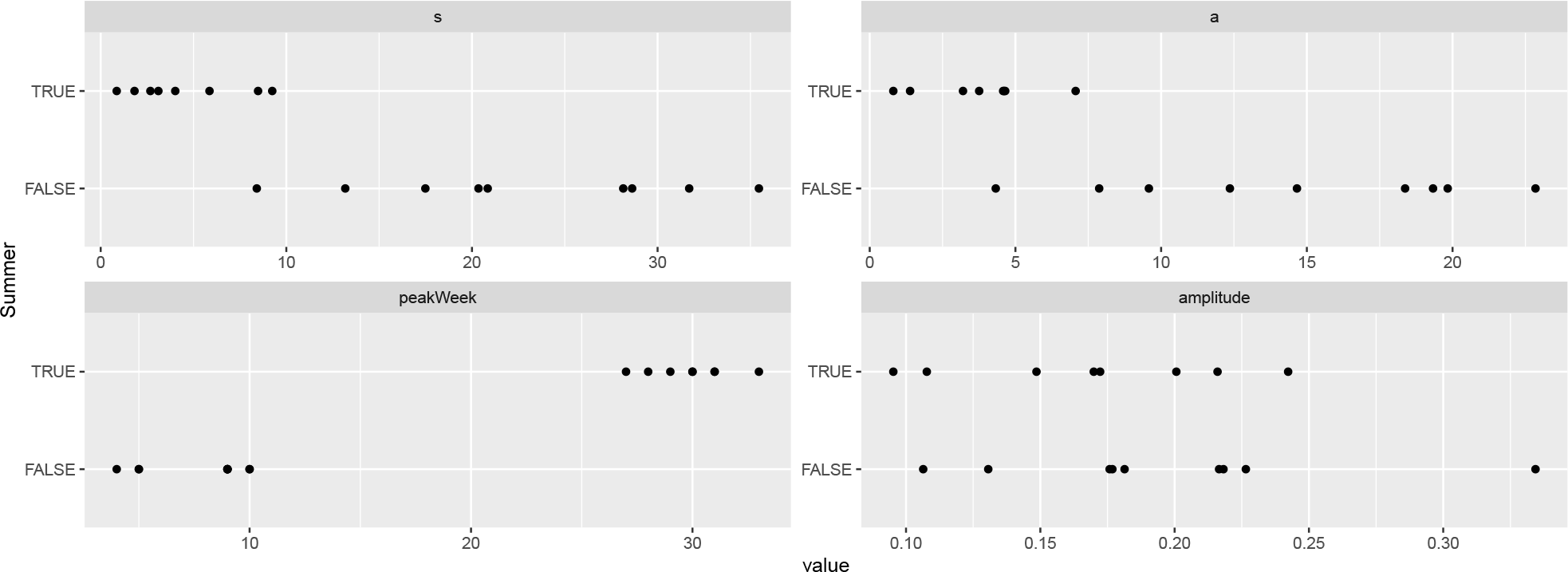
Distribution of the parameters of the best fitting rescaled Cauchy densities for each peak, separated according to whether the peak is during the summer.

This verifies that the width is indeed different, with the *s* of the summer peaks being below 10, and the winter peaks being above, i.e., summer peaks are shorter in duration, raise and fall faster. Interestingly, the peak heights are not substantially different between winter and summer. Also note that the probability of having a peak at all is different: there are 8 summer peaks and 9 winter peaks (from 20 years). Winter peaks occur between weeks 4 and 10, summer peaks occur from weeks 25 to 35.

This allows the removal of the peaks (Figure 14), and, after these peaks are removed, it is possible to re-estimate trend and seasonality, now without the biasing effect of the peaks. This “bootstrap” procedure is adequate after this second iteration, as no further peaks can be seen after the removal of the re-estimated trend and seasonality.

**Figure 14:**
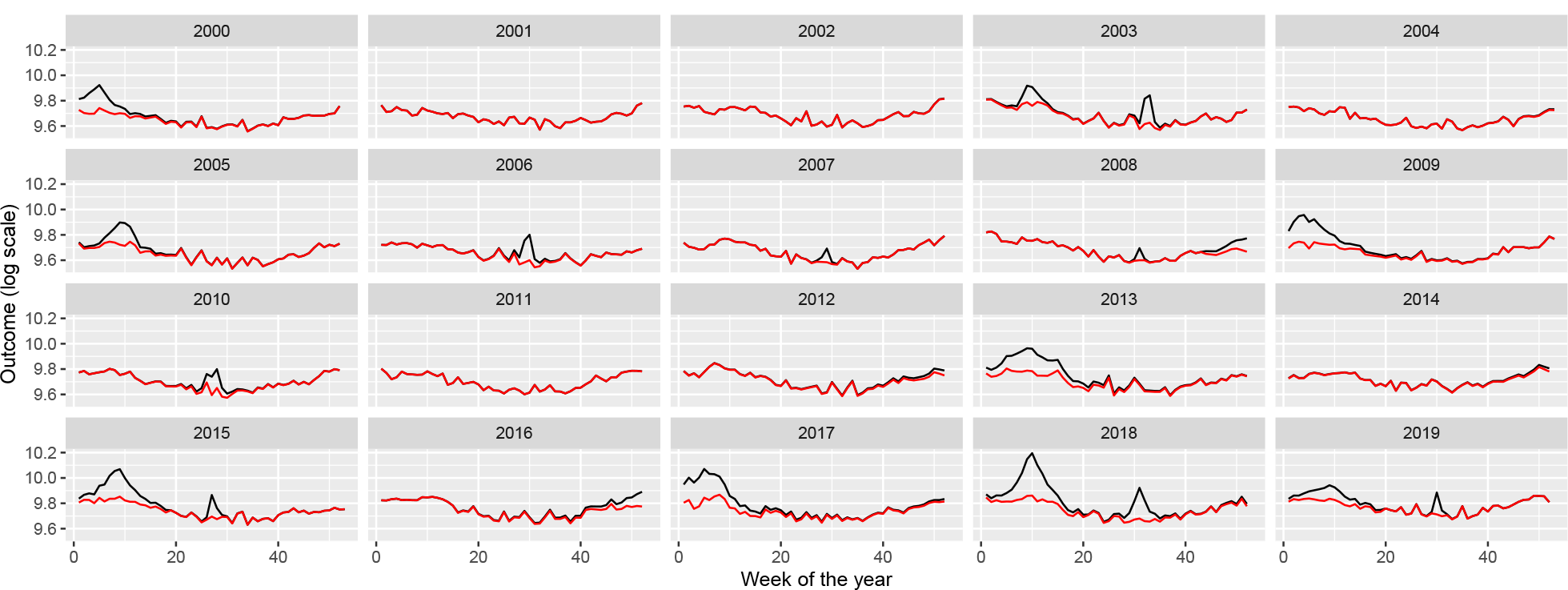
Weekly number of deaths in Germany, separated according to year (black) and with peaks removed (red).

Creating the appropriate model to simulate such peaks is not straightforward: there is stochasticity in the position of the peaks and in its shape, i.e., height and width. (Actually, even the presence of a peak is stochastic.) The following procedure will be used: the presence is generated as a Bernoulli random variate (with the probabilities described above, different for summer and winter), the onset date is uniformly distributed (from 0 to 0.2 in scaled weeks for the winter peak and from 0.5 to 0.7 for the summer peak), i.e., the position itself is random, but the parameters of the underyling distribution are fixed. The *b* parameter is set to zero (irrespectively of its estimated value, to really capture only the peak, locally – a non-zero *b* would mean a non-local effect), while *s* and *a* are randomly generated for each peak, again, separately for summer and winter peaks. Given the high correlation between *s* and *a*, not these, but rather *s* (width) and the amplitude will be generated as a random variate from – independent – uniform distributions. The parameters (minimum and maximum) of the uniform distributions both for *s* and the amplitude will be considered as a parameter (hyperparameter) of the simulational procedure, just as the *p* probability of the Bernoulli distribution, all different for summer and winter.

The following table summarizes the parameters:

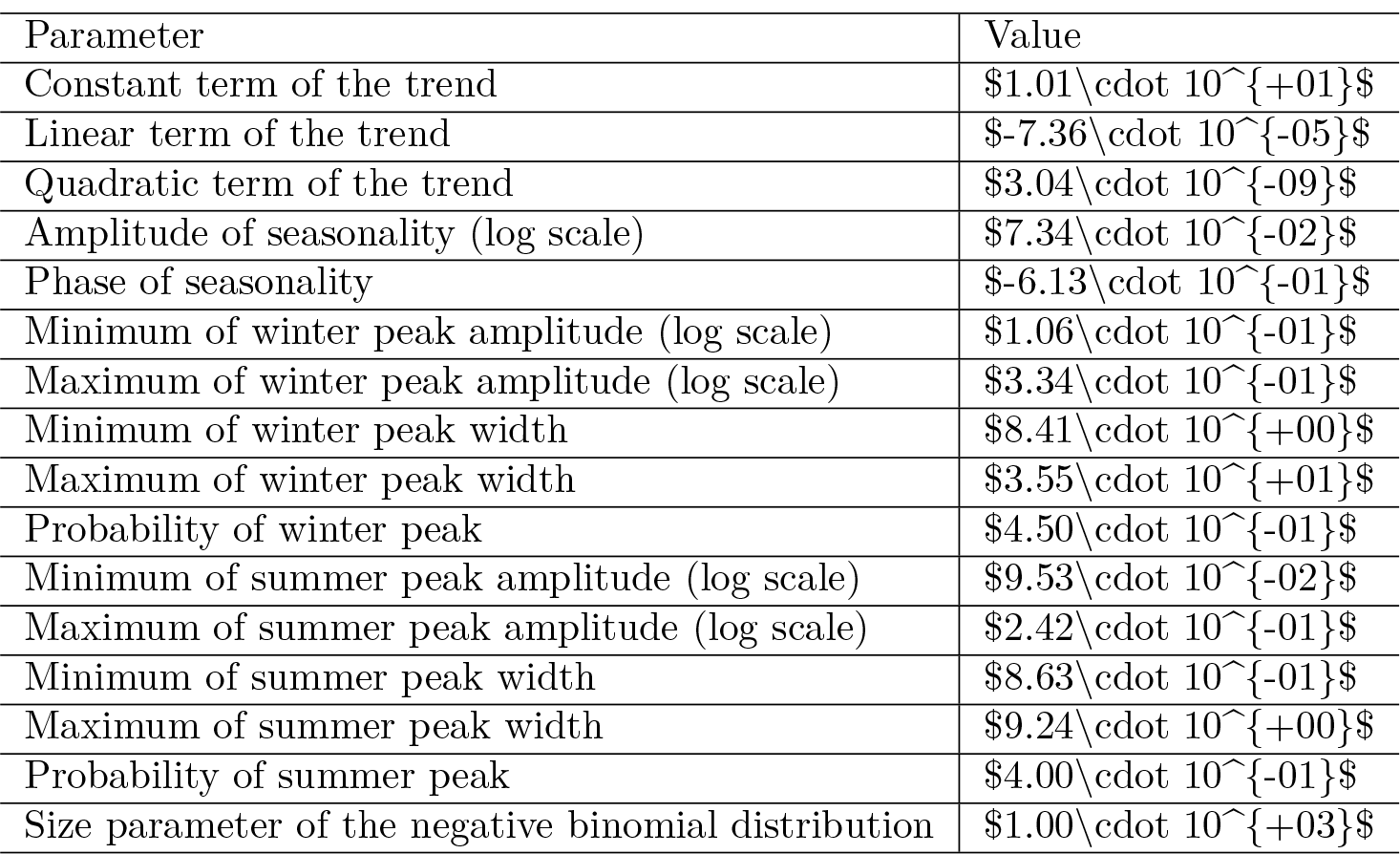

| Parameter | Value |
| --- | --- |
| Constant term of the trend | $\$1.01 \cdot 10^{+01}$ |
| Linear term of the trend | $\$-7.36 \cdot 10^{-05}$ |
| Quadratic term of the trend | $\$3.04 \cdot 10^{-09}$ |
| Amplitude of seasonality (log scale) | $\$7.34 \cdot 10^{-02}$ |
| Phase of seasonality | $\$-6.13 \cdot 10^{-01}$ |
| Minimum of winter peak amplitude (log scale) | $\$1.06 \cdot 10^{-01}$ |
| Maximum of winter peak amplitude (log scale) | $\$3.34 \cdot 10^{-01}$ |
| Minimum of winter peak width | $\$8.41 \cdot 10^{+00}$ |
| Maximum of winter peak width | $\$3.55 \cdot 10^{+01}$ |
| Probability of winter peak | $\$4.50 \cdot 10^{-01}$ |
| Minimum of summer peak amplitude (log scale) | $\$9.53 \cdot 10^{-02}$ |
| Maximum of summer peak amplitude (log scale) | $\$2.42 \cdot 10^{-01}$ |
| Minimum of summer peak width | $\$8.63 \cdot 10^{-01}$ |
| Maximum of summer peak width | $\$9.24 \cdot 10^{+00}$ |
| Probability of summer peak | $\$4.00 \cdot 10^{-01}$ |
| Size parameter of the negative binomial distribution | $\$1.00 \cdot 10^{+03}$ |

Given the mechanism described above, the procedure to simulate synthetic datasets can easily be created. Of course, as every calculation is carried out on the log scale, the mean should be exponentiated at the last step.

Figure 15 illustrates the synthetic data set creation with a single simulation. (Of course, to assess the correctness of the simulation, several realizations have to be inspected.) In addition to the plots of 2, it also gives the autocorrelation function so that it can also be compared. (The simulated outcomes are themselves independent – meaning that effects like the increased probability of a flu season if the previous year did not have one are neglected –, but the trend, seasonality and the peaks induce a correlation structure.)

**Figure 15:**
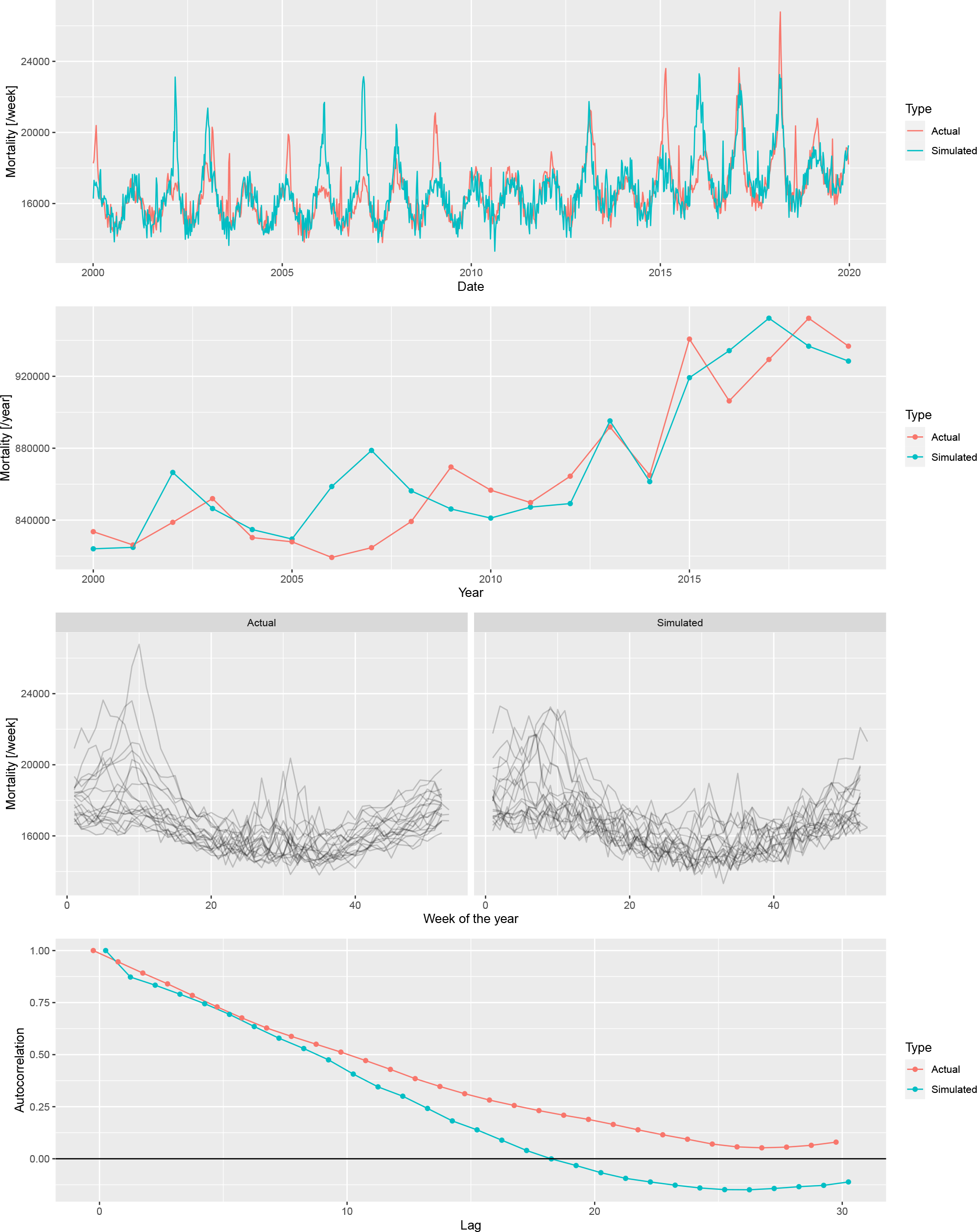
From top to bottom: weekly mortalities, yearly mortalities, seasonal pattern and autocorrelation function of the actual German mortality data and a single simulated dataset, 2000-2019.

Several simulations confirm an overall good fit between the actual data set and the simulated ones. Thus, it is now possible to investigate the properties of the mortality prediction on algorithms using simulated datasets, where the actual outcome is known, and the parameters can be varied.

## Additional File 3: Validation through simulation

Two sets of parameters have to be set up: parameters of the simulation (i.e., parameters of the scenario, on which the methods will be run) and parameters of the methods. They’re set up as given in the main text.

One thousand simulation will be run for each parameter of the scenario, and for each of the 1000 simulated data set all 4 methods with all possible parameters of the methods will be evaluated. To increase the speed, simulations will be run in parallel. (The problem is embarrassingly parallel, as different simulations are completely independent of each other [51].)

## Additional File 4: Directly comparing the errors in simulation runs

As different methods were evaluated on the same simulated dataset for each simulation, it is possible to compare not only the averages, but directly compare the errors themselves. Figure 16 shows direct comparison between the best parametrization of the WHO’s method and the Acosta-Irizarry method for 200 randomly selected simulations in each scenario, with 2015 as the starting year. In the base case scenario, Acosta-Irizarry performed better in 55.4% of the cases.

**Figure 16:**
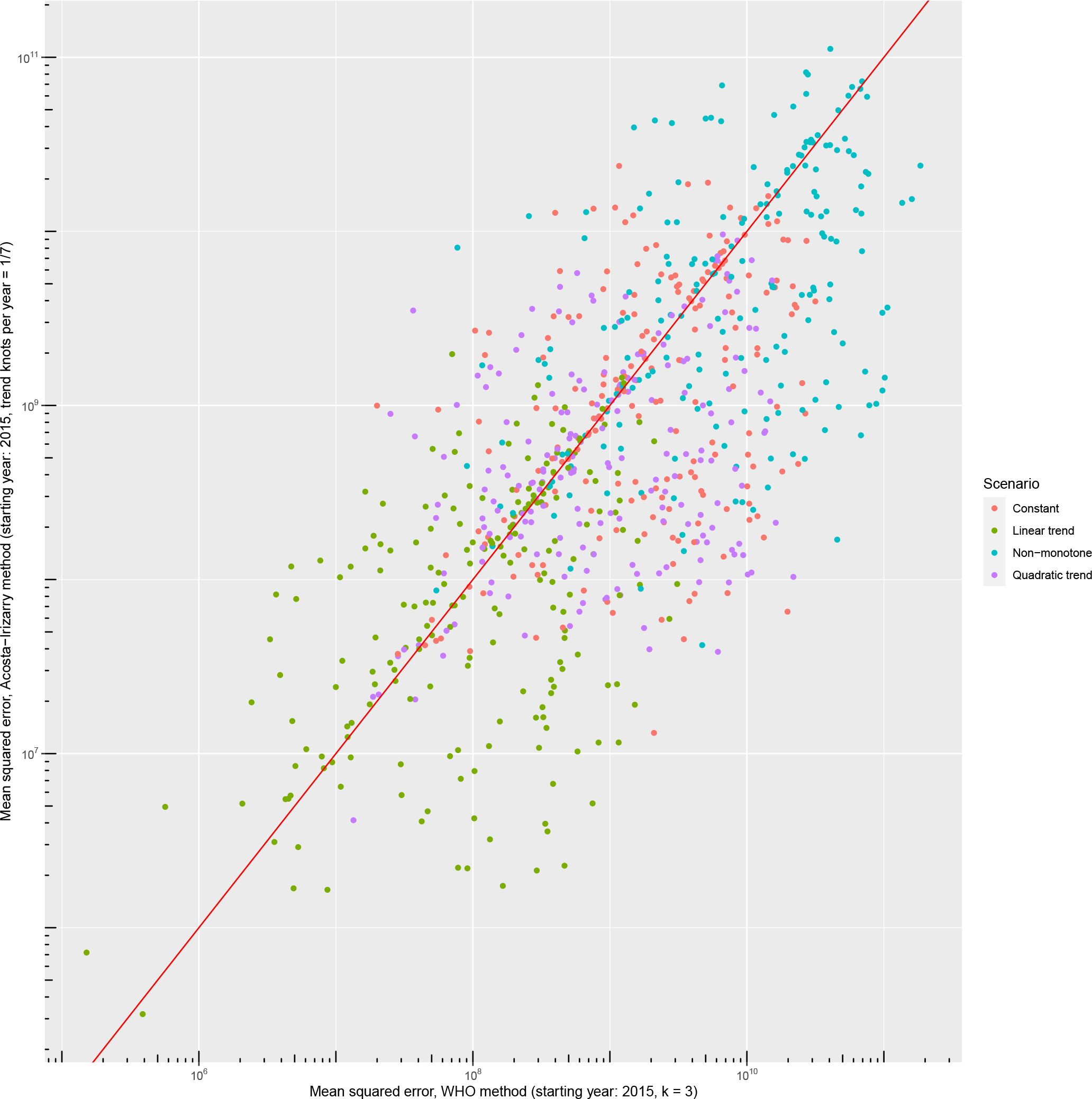
Errors – squared distance of the predicted outcome from its true value – of the WHO’s method and the Acosta-Irizarry method (under best parametrization) on the same simulated datasets for 200 randomly selected simulations with different scenarios; black dots indicate the base case scenario, scenario #1 to #5 represent varying the parameter shown on the panel from half of its base case value to twice (with the exception of the constant term where it is varied from 90% to 110%), and probabilities are limited to be below 100%.

